# Insomnia and early incident atrial fibrillation: A 16-year cohort study of younger men and women Veterans

**DOI:** 10.1101/2023.03.28.23287889

**Authors:** Allison E. Gaffey, Lindsey Rosman, Rachel Lampert, Henry K. Yaggi, Sally G. Haskell, Cynthia A. Brandt, Alan D. Enriquez, Anthony J. Mazzella, Matthew M. Burg

## Abstract

**Background:** There is growing consideration of sleep disturbances and disorders in early cardiovascular risk, including atrial fibrillation (AF). Obstructive sleep apnea (OSA) confers risk for AF but is highly comorbid with insomnia, another common sleep disorder. The objectives of this investigation were first to determine the association of insomnia and early incident AF risk and second, to determine if AF onset is earlier among those with insomnia.

**Methods:** This retrospective analysis used electronic health records from a cohort study of U.S. Veterans who were discharged from military service as of October 1, 2001 (i.e., post-9/11) and received Veterans Health Administration (VA) healthcare, 2001-2017. Time-varying, multivariate Cox proportional hazard models were used to examine the independent contribution of insomnia diagnosis to AF incidence while serially adjusting for demographics, lifestyle factors, clinical comorbidities including OSA, psychiatric disorders, and healthcare utilization.

**Results:** Overall, 1,063,723 post-9/11 Veterans (*Mage*=28.2 years, 12% women) were followed for 10 years on average. There were 4168 cases of AF (0.42/1000 person-years). Insomnia was associated with a 32% greater, adjusted risk of AF (95% CI, 1.21-1.43), and Veterans with insomnia showed AF onset up to two years earlier. Insomnia-AF associations were similar after accounting for healthcare utilization, excluding Veterans with OSA, and among those with a sleep study (adjusted hazard ratios [aHR]: 1.29-1.34).

**Conclusions:** In younger adults, insomnia was independently associated with incident AF even when accounting for OSA. Additional studies should determine if this association differs by sex and if behavioral or pharmacological treatment for insomnia attenuates AF risk.

**Clinical Perspective:** *What is new?:*

- In more than one million younger men and women Veterans with 16 years of follow-up, a history of insomnia conferred a 32% increase in risk for atrial fibrillation (AF).
- The insomnia-AF association persisted despite accounting for obstructive sleep apnea, a well-known risk factor for AF, and other demographic, lifestyle, and clinical factors.
- Veterans with insomnia may also present with AF up to 2 years earlier compared to those without insomnia.

*What are the clinical implications?:*

- Insomnia is a potentially modifiable risk factor for AF and sleep should be a focus for AF prevention.
- Screening and referral for insomnia and other sleep symptoms is critical, particularly among patients with an elevated risk for cardiovascular disease, as observed in the Veteran population.
- Although sex-specific associations could not be examined, insomnia is more prevalent among women, and it is possible that insomnia-AF associations may differ by sex.

## Introduction

Atrial fibrillation (AF) is the most prevalent sustained arrhythmia and is associated with an increased risk of major adverse cardiovascular events, related mortality, and impaired quality of life.^1^ Yet, AF typically presents after age 60.^1^ Compared to AF diagnosed later in life, the occurrence of AF in younger adults leads to a greater lifelong treatment and financial burden^2^ and a higher associated risk of mortality.^3^ Improving AF prevention and associated cardiovascular risk mitigation among younger adults requires identifying and understanding the confluence of modifiable behavioral (e.g., obesity, smoking, substance use) and cardiovascular factors (e.g., high blood pressure, diabetes) that contribute to the early onset of AF.^4-6^

In 2022 the American Heart Association added sleep as an essential component of cardiovascular health,^7^ and a recent National Heart, Lung, and Blood Institute report highlighted understanding sleep disturbances and disorders as an area of scientific priority to inform AF prevention.^8^ Obstructive sleep apnea (OSA), a prevalent sleep disorder, is prominently associated with risk for AF.^9^ Insomnia is another common sleep disorder, including among younger adults,^10,28^ defined as difficulty falling or staying asleep, which may result in daytime fatigue and dysfunction.^11^ Strong observational evidence shows that insomnia is associated with risk for various cardiovascular diseases.^12^ Insomnia may be associated with risk for AF, but studies have been limited by reliance on cross-sectional or self-report data, failing to address OSA, or only focusing on older adults.^13-17^

Veterans are more vulnerable to developing insomnia than non-Veterans,^18^ and are thereby a population of interest for understanding the potential risk associated with AF. This investigation utilized data from a nationwide cohort of 1.1 million young and middle-aged U.S. Veterans who were discharged from military service since the 9/11 terrorist attacks. The primary objective was to determine the independent association of an insomnia diagnosis to risk for incident AF. We hypothesized that insomnia would be associated with an increased risk of developing early AF after controlling for OSA and other lifestyle and clinical risk factors. We also explored if AF would occur at a younger age among those with insomnia.

## Methods

### Patient Population and Data Sources

Data were obtained from the U.S. Department of Defense’s Manpower Data Center, which includes records from individuals who served in support of post-9/11 operations and enrolled in Veterans Affairs medical center (VA) care after their discharge (*N* = 1,063,723). Records were merged with administrative, diagnostic, and pharmacy data, including outpatient encounters and inpatient hospitalizations (*International Classification of Diseases, Ninth Revision, Clinical Modification* and *Tenth Revision* [*ICD-9-CM and 10*] codes and dates), from the VA Corporate Data Warehouse.^19^ Veterans with at least 2 clinical encounters at a VA facility from 10/1/01 to 12/31/17 and no history of AF at their first VA encounter were included. EHR diagnoses were included if an ICD-9-CM or 10 code was recorded during 1 inpatient or ≥2 outpatient encounters prior to the first evidence of AF. Waiver of informed consent and procedures were approved by the VA Connecticut Healthcare System Institutional Review Board. As VA records are proprietary to the U.S. Government, the data are unavailable.

### Primary Exposure: Insomnia Diagnosis

Insomnia diagnoses were identified based on ICD-9-CM or 10 codes during follow-up (e.g., 307.x; 780.51, 780.52; F51.x; G47.x).^19^

### Outcome Ascertainment: Incident AF

The primary outcome was a new diagnosis of AF, defined as an ICD-9-CM or 10 code (e.g., 427.x, I48.x) documented in VA encounters. Consistent with VA diagnostic procedures, AF diagnoses were validated based on ≥1 inpatient or outpatient Current Procedural Terminology (CPT) code indicating assessment for AF with an electrocardiogram (93000, 93005, 93010), traditional Holter (24-48 h; 93224, 93225, 93226, 93227), extended Holter (>48 h-21 day; 0295T, 0296T, 0297T, 0298T), or an event monitor (93268, 93270, 93271, 93272, 93228, 93229) within 8 weeks before an AF diagnosis. Data concerning AF duration were unavailable, and distinctions between paroxysmal, persistent, or permanent AF could not be made.

### Other Risk Factors for AF

Other relevant variables included demographics (age, sex, race/ethnicity [Black, Hispanic, White, Other (American Indian or Alaska Native; Asian; Native Hawaiian or Other Pacific Islander, mixed race, declined, or unknown)], and lifestyle factors (obesity [body mass index (BMI) ≥30 kg/m^2^],^20^ and smoking status [current, former, never]). Validated ICD-9-CM and 10 codes were used to identify diagnoses of alcohol abuse, drug use (aggregated from use of cocaine, opioids, sedative/hypnotics, other drugs, or polysubstance use), OSA, hypertension, lipid disorders, diabetes, congestive heart failure (CHF), coronary artery disease (CAD), myocardial infarction (MI), major depressive disorder (MDD), generalized anxiety disorder (GAD), and posttraumatic stress disorder (PTSD).^21^ Given the high multicollinearity between psychiatric comorbidities,^22^ a composite variable was created from the presence of either GAD, MDD, or PTSD, with time to event based on the first diagnosis. For sensitivity analyses, CPT codes were used to identify individuals who were prescribed a continuous positive airway pressure (i.e., CPAP) device and with a history of a sleep study (e.g., 95800, 95801, 95805, 95806, G0399, G0400), to better categorize those with OSA.

Prescription data for antihypertensives (e.g., angiotensin-converting enzyme inhibitors, angiotensin receptor blockers, beta-blockers), stimulants (i.e., amphetamines, methylphenidate), and medications used to manage sleep (e.g., estazolam, eszopiclone, melatonin, trazodone) were obtained from VA pharmacy records to reflect relevant medication use. Healthcare utilization was defined as total primary care visits during the first 2 years of VA care.^19^

### Statistical Analysis

SAS V9.4 (Cary, North Carolina, USA) was used for analyses, with *p*<0.05 (two-tailed) indicating statistical significance. Baseline was the date of a Veteran’s first VA clinical encounter. Follow-up time extended from the date of the first clinical encounter until whichever occurred first: an AF diagnosis or the date of data censoring (last VA encounter) and was assessed individually. After Veterans met criteria for AF, further diagnoses and prescriptions were excluded.

Machine-learning clinical algorithms that include objective variables (e.g., age, sex, BMI, comorbidities) can better predict OSA than subjective data from questionnaires. One such algorithm developed by Ustun et al.,^23^ was used to determine patients who might have undiagnosed OSA. Algorithm scores were calculated based on age, BMI, diabetes, hypertension, smoking status, and female sex category. Individuals who scored >29 were predicted to have OSA and were included in the OSA group in secondary analyses, whereas the primary analyses only used those with ICD-9-CM and 10 diagnoses of OSA. Cumulative incidence rates of AF per 1,000 person-years were calculated. Kaplan-Meier cumulative incidence functions were used to display AF incidence across follow-up, stratified by the presence of an insomnia diagnosis.

Time-varying Cox proportional-hazards models with 95% confidence intervals (CIs) were used to evaluate the unadjusted independent association of insomnia and risk for AF. Four additional multivariate models then sequentially adjusted for the following *a priori* covariates: 1) demographics (age, sex [female as reference], race/ethnicity [White as reference]); 2) modifiable lifestyle and health factors (BMI, smoking status [“Never smoked” as reference], illicit drug use, and alcohol abuse); 3) clinical risk factors (e.g., OSA, hypertension), and 4) psychiatric comorbidities (GAD, MDD, and PTSD). Given the time-dependent associations between insomnia, AF, and other diagnoses (e.g., diabetes, MDD), these variables were modeled as time-varying, with each variable coded as 0 prior to, and 1 following, the diagnosis date. We excluded clinical diagnoses after the index AF event. Log-rank tests were used to determine if the time to event differed by an insomnia diagnosis. Finally, sensitivity analyses assessed for surveillance bias based on healthcare utilization, OSA, and sleep study participation.

To compensate for missing data (i.e., BMI: 22%, healthcare utilization: 12%, smoking status: 26%), regression-based multiple imputation was used to generate 5 data sets with complete values. Analysis of Schoenfeld residuals indicated no violation of the proportional hazards assumption.

## Results

Table 1 illustrates the sample of 1,063,723 Veterans, with characteristics presented overall and by insomnia diagnosis. The average age at baseline was 28.2 (SD:8.8) years. Most Veterans were male (87%), White (65%), and non-Hispanic (89%), almost half were married (45%), and a majority had served in active duty (61%). Approximately 11% of the cohort were diagnosed with insomnia, a quarter were categorized as obese (26%), and more than half (60%) were past or current smokers. The most prevalent clinical and psychiatric diagnoses were lipid disorders (18%) and PTSD (27%). About 6% of the cohort had undergone a sleep study.

**Table 1.**
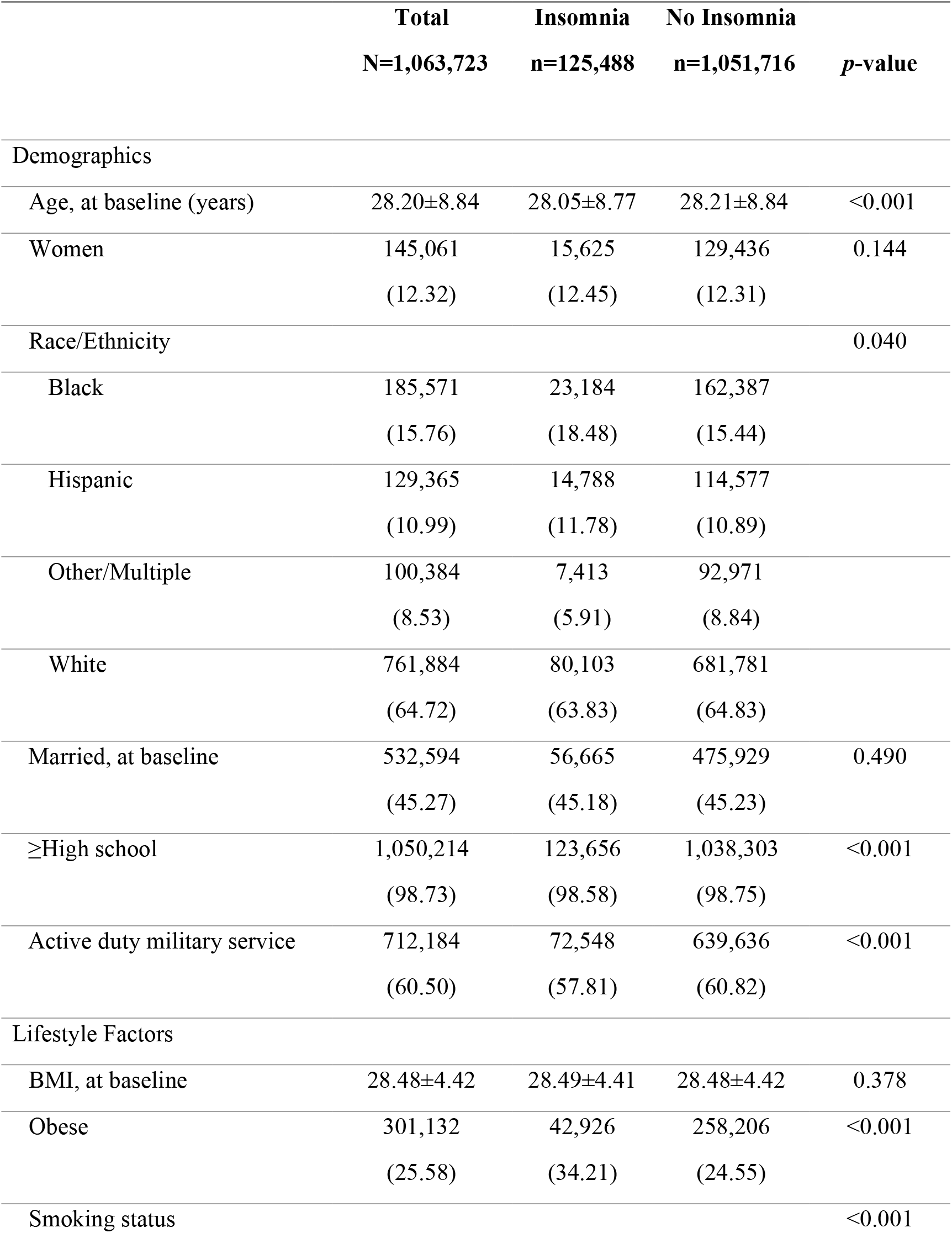
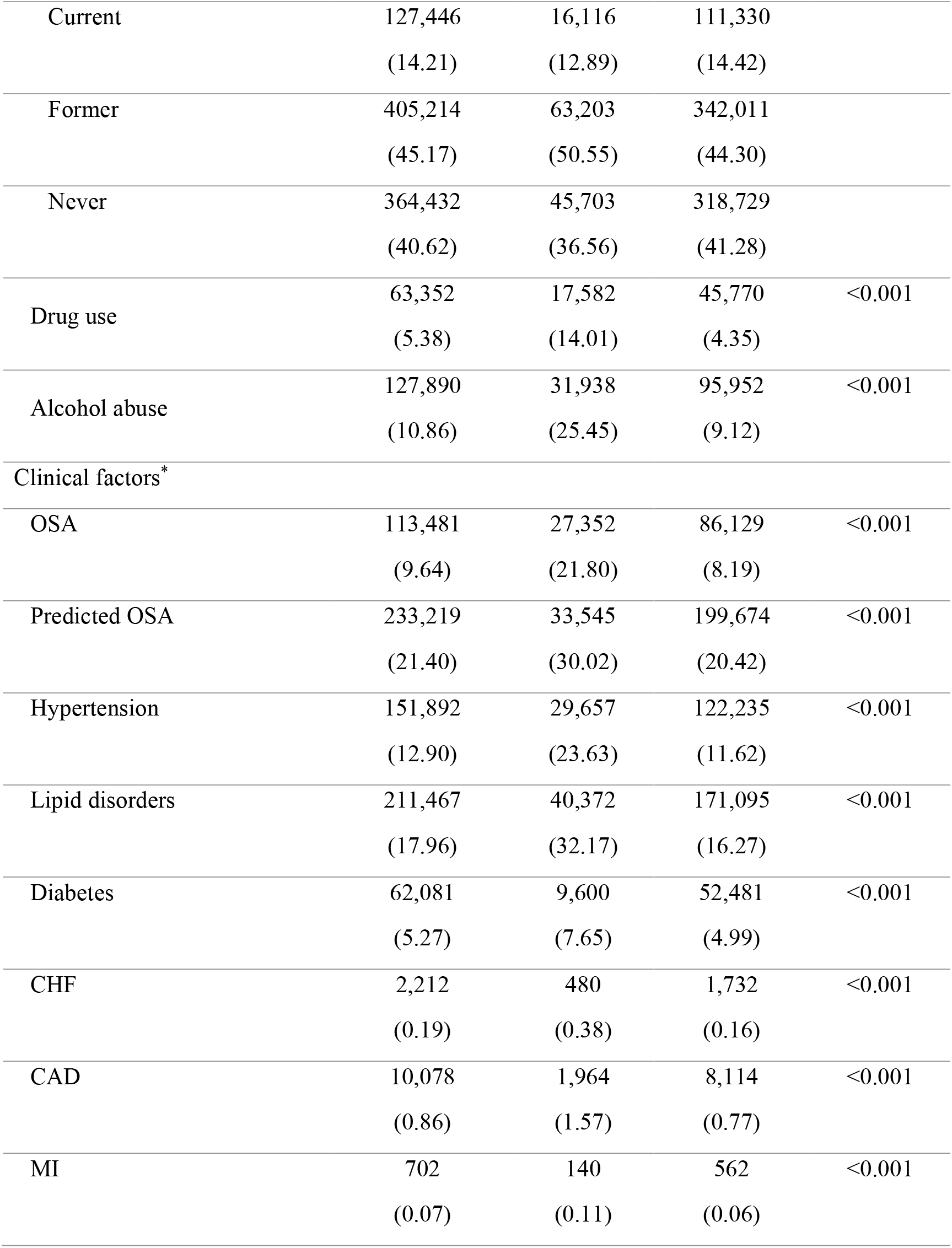
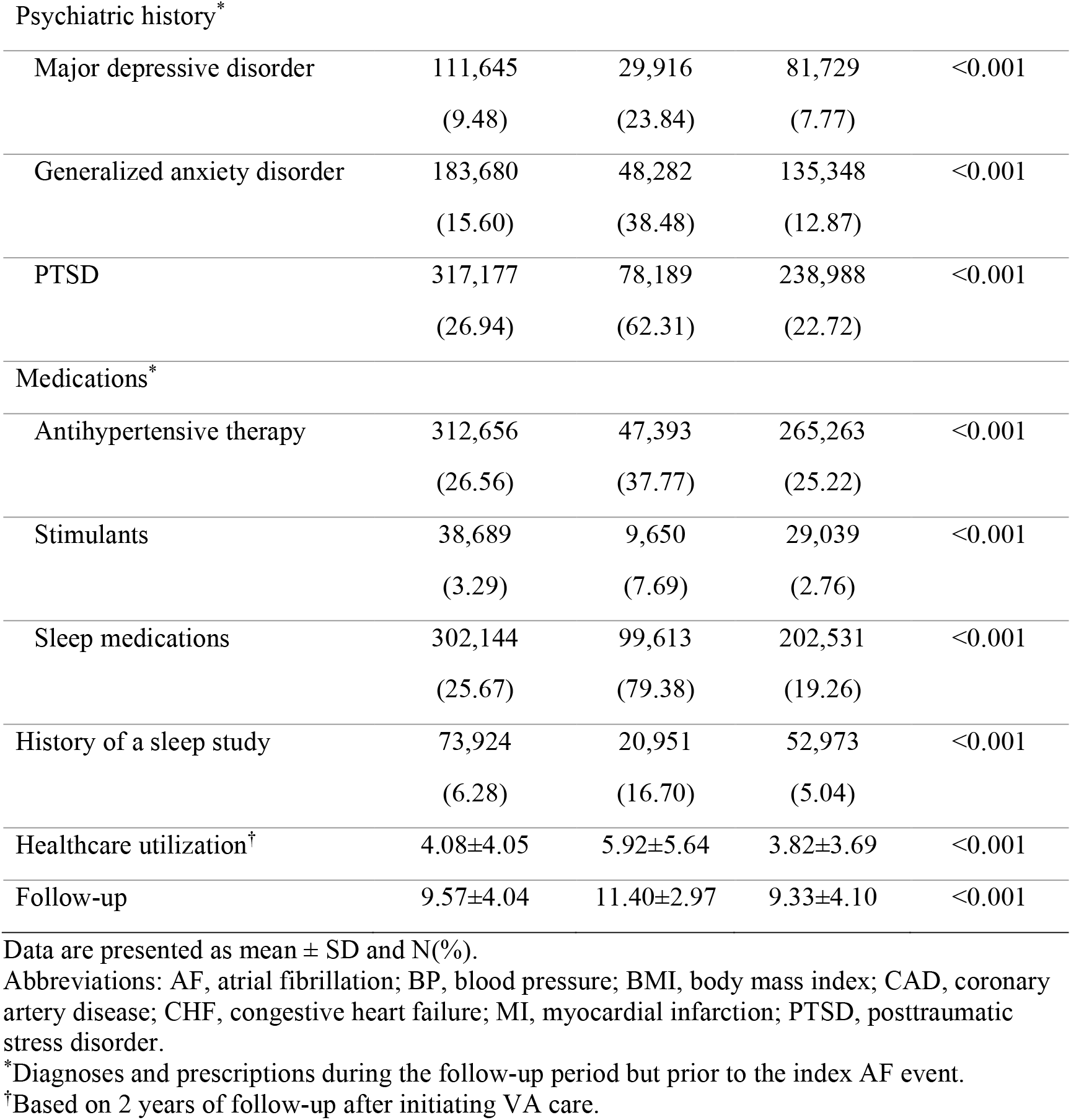
Characteristics of the Cohort, Overall and by Insomnia Diagnosis.

|  | **Total**<br>**N=1,063,723** | **Insomnia**<br>**n=125,488** | **No Insomnia**<br>**n=1,051,716** | ***p*-value** |
| --- | --- | --- | --- | --- |
| **Demographics** |  |  |  |  |
| Age, at baseline (years) | 28.20±8.84 | 28.05±8.77 | 28.21±8.84 | <0.001 |
| Women | 145,061<br>(12.32) | 15,625<br>(12.45) | 129,436<br>(12.31) | 0.144 |
| Race/Ethnicity |  |  |  | 0.040 |
| Black | 185,571<br>(15.76) | 23,184<br>(18.48) | 162,387<br>(15.44) |  |
| Hispanic | 129,365<br>(10.99) | 14,788<br>(11.78) | 114,577<br>(10.89) |  |
| Other/Multiple | 100,384<br>(8.53) | 7,413<br>(5.91) | 92,971<br>(8.84) |  |
| White | 761,884<br>(64.72) | 80,103<br>(63.83) | 681,781<br>(64.83) |  |
| Married, at baseline | 532,594<br>(45.27) | 56,665<br>(45.18) | 475,929<br>(45.23) | 0.490 |
| ≥High school | 1,050,214<br>(98.73) | 123,656<br>(98.58) | 1,038,303<br>(98.75) | <0.001 |
| Active duty military service | 712,184<br>(60.50) | 72,548<br>(57.81) | 639,636<br>(60.82) | <0.001 |
| **Lifestyle Factors** |  |  |  |  |
| BMI, at baseline | 28.48±4.42 | 28.49±4.41 | 28.48±4.42 | 0.378 |
| Obese | 301,132<br>(25.58) | 42,926<br>(34.21) | 258,206<br>(24.55) | <0.001 |
| Smoking status |  |  |  | <0.001 |
| Current | 127,446<br>(14.21) | 16,116<br>(12.89) | 111,330<br>(14.42) |  |
| Former | 405,214<br>(45.17) | 63,203<br>(50.55) | 342,011<br>(44.30) |  |
| Never | 364,432<br>(40.62) | 45,703<br>(36.56) | 318,729<br>(41.28) |  |
| Drug use | 63,352<br>(5.38) | 17,582<br>(14.01) | 45,770<br>(4.35) | <0.001 |
| Alcohol abuse | 127,890<br>(10.86) | 31,938<br>(25.45) | 95,952<br>(9.12) | <0.001 |
| Clinical factors* |  |  |  |  |
| OSA | 113,481<br>(9.64) | 27,352<br>(21.80) | 86,129<br>(8.19) | <0.001 |
| Predicted OSA | 233,219<br>(21.40) | 33,545<br>(30.02) | 199,674<br>(20.42) | <0.001 |
| Hypertension | 151,892<br>(12.90) | 29,657<br>(23.63) | 122,235<br>(11.62) | <0.001 |
| Lipid disorders | 211,467<br>(17.96) | 40,372<br>(32.17) | 171,095<br>(16.27) | <0.001 |
| Diabetes | 62,081<br>(5.27) | 9,600<br>(7.65) | 52,481<br>(4.99) | <0.001 |
| CHF | 2,212<br>(0.19) | 480<br>(0.38) | 1,732<br>(0.16) | <0.001 |
| CAD | 10,078<br>(0.86) | 1,964<br>(1.57) | 8,114<br>(0.77) | <0.001 |
| MI | 702<br>(0.07) | 140<br>(0.11) | 562<br>(0.06) | <0.001 |
| Psychiatric history* |  |  |  |  |
| Major depressive disorder | 111,645<br>(9.48) | 29,916<br>(23.84) | 81,729<br>(7.77) | <0.001 |
| Generalized anxiety disorder | 183,680<br>(15.60) | 48,282<br>(38.48) | 135,348<br>(12.87) | <0.001 |
| PTSD | 317,177<br>(26.94) | 78,189<br>(62.31) | 238,988<br>(22.72) | <0.001 |
| Medications* |  |  |  |  |
| Antihypertensive therapy | 312,656<br>(26.56) | 47,393<br>(37.77) | 265,263<br>(25.22) | <0.001 |
| Stimulants | 38,689<br>(3.29) | 9,650<br>(7.69) | 29,039<br>(2.76) | <0.001 |
| Sleep medications | 302,144<br>(25.67) | 99,613<br>(79.38) | 202,531<br>(19.26) | <0.001 |
| History of a sleep study | 73,924<br>(6.28) | 20,951<br>(16.70) | 52,973<br>(5.04) | <0.001 |
| Healthcare utilization† | 4.08±4.05 | 5.92±5.64 | 3.82±3.69 | <0.001 |
| Follow-up | 9.57±4.04 | 11.40±2.97 | 9.33±4.10 | <0.001 |

Data are presented as mean ± SD and N(%).
Abbreviations: AF, atrial fibrillation; BP, blood pressure; BMI, body mass index; CAD, coronary artery disease; CHF, congestive heart failure; MI, myocardial infarction; PTSD, posttraumatic stress disorder.
\*Diagnoses and prescriptions during the follow-up period but prior to the index AF event.
†Based on 2 years of follow-up after initiating VA care.

An OSA diagnosis was recorded for 10% of Veterans, and there was a significantly higher prevalence of OSA among those with an insomnia diagnosis than those without. Based on the clinical prediction algorithm,^23^ 15% of those without an OSA diagnosis in the EHR were likely to have OSA. When comparing the groups of those with diagnosed OSA versus predicted OSA, the clinical prediction tool had a sensitivity of 80.4%, and a specificity of 51.6%.

Overall, there were 4168 cases of AF (4.1% were observed in women). The likelihood of AF was 0.42 events/1,000-person-years. For those with an insomnia diagnosis, the likelihood of AF was 0.57 compared to 0.39 for those without a diagnosis (Figure 1). For men with insomnia the prevalence of AF was 0.53% and for women with insomnia the prevalence of AF was 0.18%.

**Figure 1.**
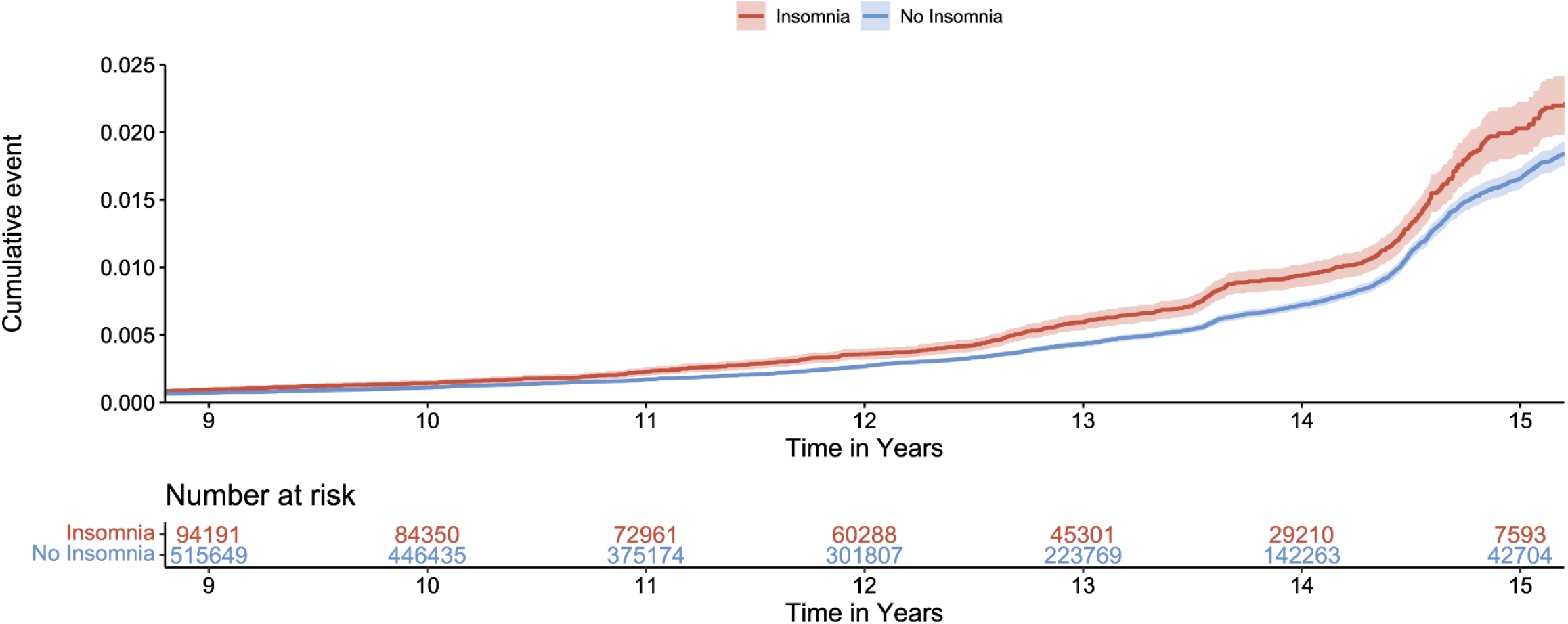
Cumulative Incidence of Atrial Fibrillation (AF) Among Younger Adults by Insomnia Diagnosis. Among a cohort of 1,063,723 post-9/11 Veterans, the cumulative incidence of AF and number of Veterans at risk, is displayed according to the presence and absence of an insomnia diagnosis. Due to a low AF incidence across the earlier years of follow-up, data is depicted as of Year 9.

In the unadjusted Cox model, individuals with insomnia showed a 62% greater risk of incident AF versus those without insomnia (95% CI, 1.50-1.74; *p*<0.001; Table 2). The insomnia-AF association was robust to adjustment for demographics, lifestyle factors, clinical factors including OSA, and psychiatric diagnoses (adjusted hazard ratio [aHR], 1.32; 95% CI, 1.21-1.43; *p*<0.001). Those with insomnia were also more likely to be diagnosed with alcohol abuse, OSA, hypertension, diabetes, CAD, CHF, and a psychiatric disorder during follow-up. AF was diagnosed at a slightly earlier age among those with insomnia compared to those without (45.09 years vs. 46.14 years, *F*(1,4165)=4.74, *p*=0.030). After excluding anyone with an OSA diagnosis, those with a history of insomnia again developed AF at a younger age (42.8 versus 45.1 years; *F*(1,2662)=12.14, *p*<0.001).

**Table 2.**
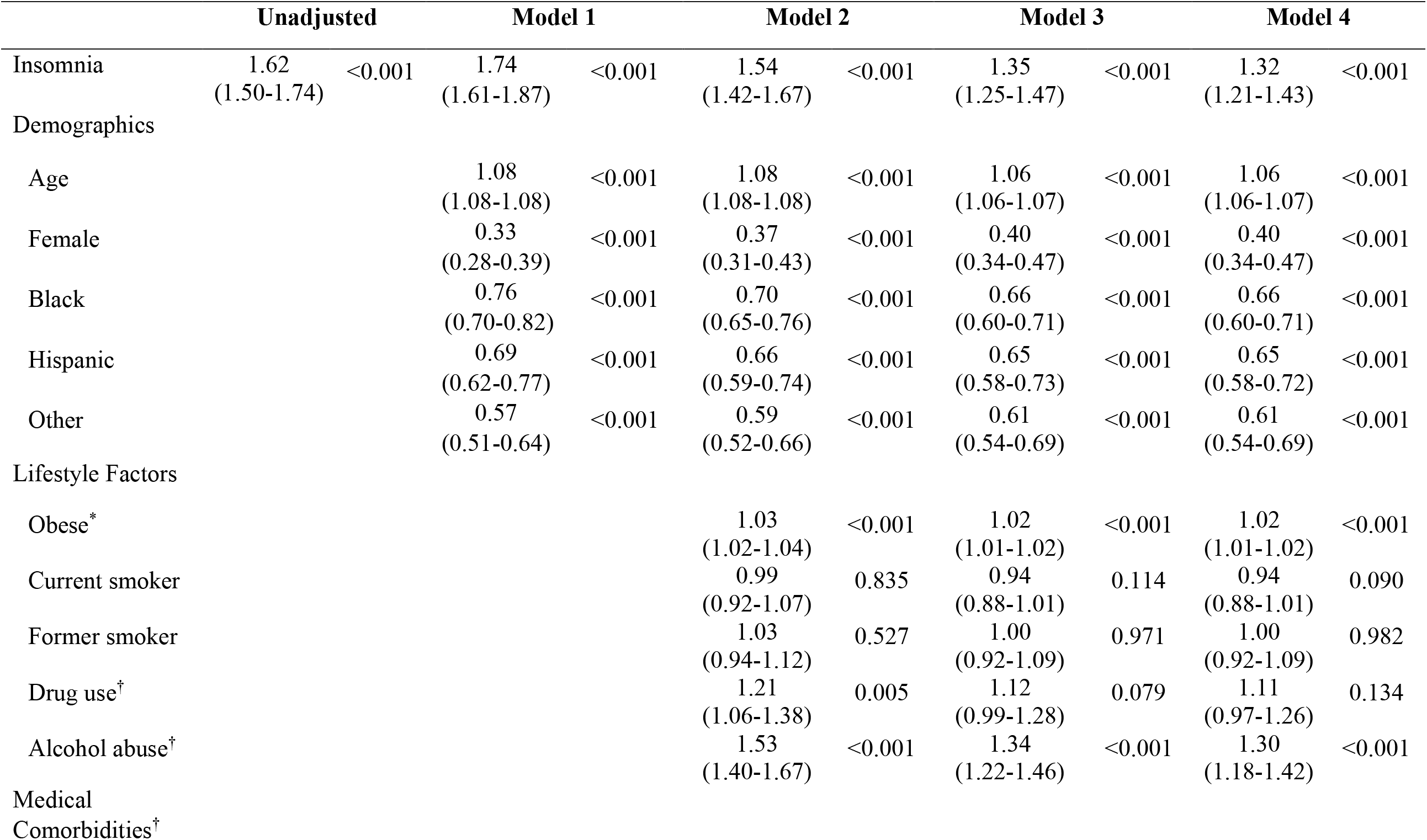
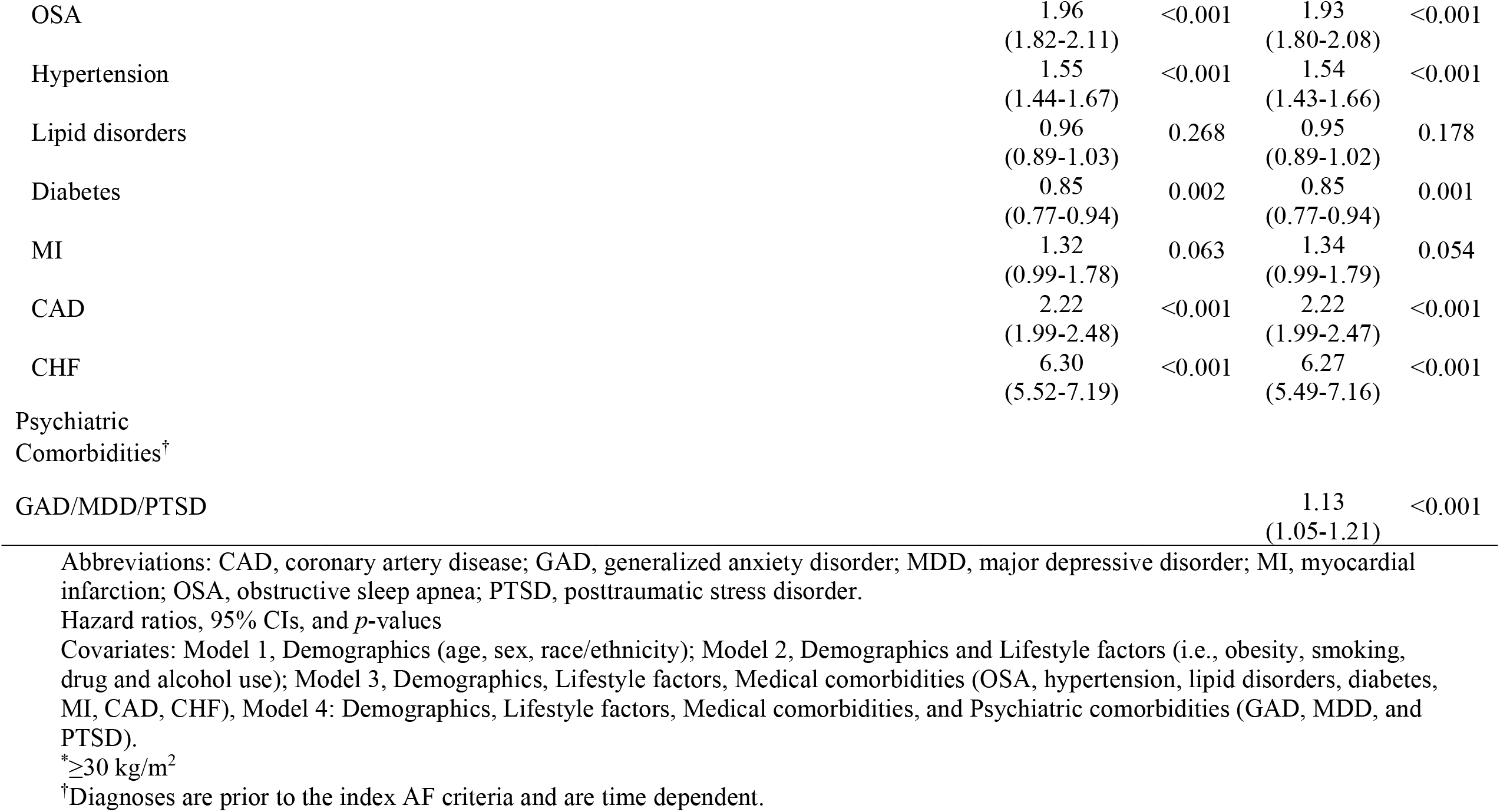
Unadjusted and Multivariate Models of Insomnia and Risk for Incident Atrial Fibrillation Insomnia Diagnosis.

The primary analyses were repeated in five sensitivity analyses. In each case, the significant, independent association of insomnia remained. First, healthcare utilization was added to the fully adjusted model. Although there was a significant association of primary care utilization on AF risk (aHR, 1.01; 95% CI, 1.00-1.30), the insomnia-AF association was robust to adjustment (aHR, 1.27; 95% CI, 1.17-1.39). Next, to further address the possibility of an association of OSA, individuals with an OSA diagnosis or with predicted OSA based on the clinical algorithm were excluded from the analytic sample (*Supplemental Tables S1 and S2*). Next, only investigating adults who had completed a sleep study (*n*=73,924), including those with diagnosed or predicted OSA (*Supplemental Table S3*). Analyses were also repeated for individuals who participated in a sleep study but who were unlikely to have OSA (diagnosed or predicted; *Supplemental Table S4*).

## Discussion

Insomnia was diagnosed in 11% of this nationwide cohort of younger military Veterans, and those with an insomnia diagnosis showed a 32% greater risk of incident AF, supporting our hypothesis. Age, male sex, BMI, a history of alcohol abuse, OSA, hypertension, CAD, CHF, and a psychiatric disorder were also associated with significantly increased AF risk, but the insomnia-AF association remained after controlling for these factors. Adults with insomnia show a higher prevalence of AF,^24^ and there are cross-sectional and prospective associations between insomnia and AF.^6,13,17,25^ But past studies were limited to substantially older samples (e.g., mean ages of 49 and ≥60 at baseline),^17,24^ did not include validated cases of AF,^6,17,24,25^ or adjust for BMI,^17^ OSA,^13^ smoking or alcohol abuse.^6^ The most compelling investigation of insomnia and AF was based on EHR data from 14 million California residents with an average age of 49 years, in which an insomnia diagnosis was associated with a 36% greater risk of incident AF over 4 years.^17^ Yet, the California sample was considerably older than our cohort. In addition to replicating insomnia-AF associations in younger adults, we demonstrated that individuals with insomnia presented with AF at a younger age than those without an insomnia diagnosis.

With considerable comorbidity between insomnia and OSA,^26^ a strength of the present investigation was the rigorous methods to exclude the possibility that the insomnia association did not simply reflect OSA, a known risk factor for AF.^9^ OSA was first included as a covariate in the primary analyses to perform traditional statistical adjustment. The primary analyses were repeated after excluding individuals with an OSA diagnosis, and also after excluding those with likely undiagnosed OSA, based on a clinical prediction algorithm.^23^ In each case, the significant association of insomnia was preserved. Although past analyses concerning insomnia and risk of incident AF also attempted to control for OSA,^6,17,25^ those studies did not distill individuals with diagnosed, treated, and *likely* yet undiagnosed OSA. Efforts to carefully rule out additional adverse sleep phenomenon and diagnoses are recommended for subsequent work to determine the unique role of insomnia in early cardiovascular risk.

Additional dimensions of disturbed sleep – e.g., sleep quality and duration – are also associated with the risk of AF and other arrhythmias.^14-17,25,27,28^ Most prominently, sleep duration – either shorter or longer than the recommended 7-8 hours – is associated with incident AF.^28-30^ Risk for incident AF may also vary based on the combination of adverse sleep symptoms. One meta-analysis of 14 million patients showed that self-report of *both* insomnia and frequent nighttime awakening was associated with a 30% higher risk of AF.^25^ Further, there are well-known age-specific patterns of insomnia – i.e., younger adults are more likely to show difficulty with sleep initiation than with nighttime wakefulness.^31^ Although data concerning specific insomnia symptoms were unavailable, and other aspects of sleep (e.g., chronotype and daytime fatigue) and the associations of insomnia were not assessed, such data could help to better characterize subgroups who are more (or less) vulnerable to AF.^16^ Finally, night-to-night variability – in sleep timing, duration, and efficiency – is another important aspect of sleep health and cardiovascular risk,^32,33^ that should be investigated in relation to AF and other arrhythmias.

### Clinical Implications

The high prevalence of insomnia in younger adults – 1 in 5^10,34^ – and the impact of AF on quality of life, medical morbidity and mortality, emphasize the importance of these findings for identifying sleep as a potentially modifiable risk factor. Specifically, the association of insomnia to AF risk was greater than those of other known AF risk factors (e.g., obesity, alcohol abuse),^35,36^ and conditions that are highly comorbid with insomnia (i.e., MDD, GAD, PTSD),^37^ suggesting that sleep should be a focus for AF prevention. Evidence supports the benefits of cognitive behavioral therapy for insomnia (CBT-I) that is comorbid with OSA and psychiatric conditions.^38-41^ CBT-I is also efficacious when offered to adults with a high risk for cardiovascular disease.^42-45^ Whether treating insomnia – with medication, CBT-I, or a combination – influences risk for AF is an important future focus.^46^

### Potential Pathophysiological Mechanisms Between Insomnia and AF

The pathophysiology linking insomnia to AF risk is not well described. One explanation relates to the potential role of psychosocial stress. For example, stress and negative emotions reported via daily diary were associated with a 2.5-fold increased likelihood of an AF episode captured with 24-h Holter monitoring.^47^ Stress could be a confounder that is associated with both insomnia and AF, or a precursor to further stress, autonomic nervous system dysfunction, and AF.^48^ Another hypothesis is that sleep fragmentation and hyperarousal, which are primary characteristics of insomnia,^11^ affect autonomic nervous system regulation (e.g., reducing heart rate variability),^49,50^ contributing to arrhythmia risk.^51^ In a recent study of young adults, individuals with insomnia, including frequent nighttime waking, displayed a lower wake-to-sleep reduction in heart rate than those without insomnia.^52^ Insomnia-associated alterations in the renin-aldosterone-angiotensin system, which is promotes atrial remodeling, have also been proposed,^53,54^ along with inflammation, endocrinological and/or adrenergic dysfunction,^55^ all of which are well-described potential mediators of AF risk.^56-58^ Other data from the UK Biobank showed that genetically-predicted insomnia is associated with a 13% higher risk of AF, and risk for eight other cardiovascular diseases.^59^ Finally, hypothalamic-pituitary-adrenal axis activity may also be involved in the physiologic pathways that are impacted by chronic sleep disruption but remains understudied.^60^ Testing of multi-system physiological markers is likely required to better explain the complex, downstream associations of insomnia on atrial rhythm.

### Strengths and Limitations

Strengths of this investigation include the cohort characteristics (e.g., a large, nationwide sample with a younger average age), the 10-year average duration of follow-up, and the ability to analytically control for diagnoses that are frequently comorbid with insomnia (e.g., OSA, PTSD). There are also limitations. First, this cohort consisted of primarily younger, male, post-9/11 Veterans and results may not generalize to older Veterans, or adults without a history of military service. Also related to generalizability, insomnia is more prevalent among younger women than men,^10^ a difference observed in middle-aged and older adults as well.^61^ Sex differences in the association between insomnia and incident AF are plausible,^62^ but with only 4% of AF cases diagnosed among women, there was insufficient statistical power to investigate this question. Exploring sex differences and sex-specific associations of insomnia and AF associations is a major future direction. Second, using administrative data to capture diagnoses of insomnia and incident AF involves selecting for subsets of patients who present to the health system and whose diagnoses are identifiable. Regrettably, insomnia is likely drastically underdiagnosed, including among younger Veterans.^63^ Similarly, there are patients with minimal AF burden who are highly symptomatic, while others have a high burden of AF (or persistent AF) who are asymptomatic. In all cases, diagnostic codes may identify the patients who are most likely to be symptomatic. Third, a propensity toward somatization or sensitivity to physical symptoms could be a potential confounding factor,^64^ and the true association between insomnia to AF may be more robust. Fourth, classifying OSA with the clinical prediction algorithm involves the potential for misclassification bias, but the algorithm showed acceptable validity for our objectives.^23^ Finally, prescriptions that were obtained outside of the VA system were unavailable and thus, not accounted for.

## Conclusions

Across 16 years, an insomnia diagnosis was associated with over a 30% greater likelihood of AF in a younger adult sample, exceeding the risk associated with other known behavioral and clinical risk factors for AF, sleep and psychiatric conditions that are comorbid with insomnia, and even after removing potential impacts of OSA. These findings suggest that insomnia is a potentially modifiable risk factor for AF, pointing the way toward intervention trials to decrease the incidence of this disease.

## Data Availability

As Department of Veterans Affairs records are proprietary to the U.S. Government, the data are unavailable.

## Nonstandard Abbreviations and Acronyms

AF: atrial fibrillation
aHR: adjusted hazard ratio
CAD: coronary artery disease
CBT-I: cognitive behavioral therapy for insomnia
CHF: congestive heart failure
CPAP: continuous positive airway pressure
GAD: generalized anxiety disorder
ICD-9-CM and 10: International Classification of Diseases, Ninth Revision, Clinical Modification and Tenth Revision
MDD: major depressive disorder
MI: myocardial infarction
OSA: obstructive sleep apnea
PTSD: posttraumatic stress disorder
VA: Veterans Affairs medical center

## Acknowledgements

The authors are grateful to all Veterans for their service and sacrifices, and especially thank the men and women who participated in the Women Veterans Cohort Study for their contribution to this work.

## Disclaimer

The views expressed in this manuscript are those of the authors and do not necessarily represent the views of the National Heart, Lung, and Blood Institute; the National Institutes of Health; the U.S. Department of Health and Human Services; or the U.S. Department of Veterans Affairs.

## Conflict of Interest Disclosures

Dr. Rosman is a consultant with Pfizer and a member of the Biotronik medical advisory board. All other authors have reported that they have no relationships relevant to the contents of this paper to disclose.

## Supplemental Material

Tables S1-S4

## Notes

### Competing Interest Statement

The authors have declared no competing interest.

### Funding Statement

This study was supported by grants from the U.S. Department of Veterans Affairs to Dr. Gaffey (VISN1 Career Development Award) and to Drs. Haskell and Brandt (IIR 12-118). Dr. Gaffey was also supported by a grant to Dr. Burg from the National Heart, Lung, and Blood Institute (R01 HL125587).

### Author Declarations

Waiver of informed consent and procedures were approved by the VA Connecticut Healthcare System Institutional Review Board.

